# Higher exposure to secondhand smoking among adolescents living in Zambia in 2021

**DOI:** 10.64898/2026.02.20.26346759

**Authors:** Wingston Felix Ng’ambi, Samuel Mutasha, Shadreck Habbanti, Adoration Chigere, Cosmas Zyambo

## Abstract

**Background:** Secondhand smoke (SHS) exposure remains a major public health concern among adolescents, particularly in low- and middle-income countries. Evidence from Zambia is limited, despite increasing tobacco use and existing tobacco control policies. This study examined the prevalence and correlates of SHS exposure among adolescents in Zambia.

**Methods:** We analyzed data from the 2021 Zambia Global Youth Tobacco Survey (GYTS), a nationally representative, school-based survey. The sample included 6,499 adolescents aged 11–17 years enrolled in grades 7–9. The primary outcome was any SHS exposure, defined as exposure to tobacco smoke at home, school, enclosed public places, or outdoor public places. Weighted prevalence estimates were calculated, and multivariable logistic regression was used to identify factors associated with SHS exposure, adjusting for demographic, social, environmental, and socioeconomic variables.

**Results:** Overall, 66.0% of adolescents reported exposure to SHS. Adolescents living with a parent or guardian who smoked had nearly three times higher odds of SHS exposure (adjusted odds ratio [AOR] = 2.76; 95% CI: 2.12–3.62; p < 0.001). Having friends who smoked tobacco (AOR = 1.86; 95% CI: 1.52–2.30; p < 0.001) and seeing teachers smoking at school (AOR = 1.88; 95% CI: 1.40–2.56; p < 0.001) were also significant predictors. Media exposure was important: seeing people use tobacco on television (AOR = 1.88; 95% CI: 1.63–2.17; p < 0.001) and exposure to tobacco advertisements (AOR = 1.38; 95% CI: 1.14–1.67; p = 0.001) increased odds of SHS exposure. Adolescents who had smoked cigarettes had higher odds of exposure (AOR = 2.80; 95% CI: 1.70–4.67; p < 0.001), as did those intending to use tobacco in the next five years (AOR = 1.64; 95% CI: 1.21–2.24; p = 0.002). Age, sex, and grade level were not independently associated with SHS exposure.

**Conclusions:** SHS exposure among adolescents in Zambia is widespread and is largely driven by household smoking, peer influence, school environments, and media exposure. Strengthening enforcement of smoke-free policies, promoting smoke-free homes, and addressing social and media influences are critical to reducing adolescent SHS exposure.

## Introduction

Second-hand smoke (SHS), also known as passive or involuntary smoking, remains a major global health concern despite declines in active smoking in many regions. Tobacco smoke contains thousands of chemicals, including numerous carcinogens and cardio-respiratory toxins, and SHS exposure has been causally linked to ischemic heart disease, stroke, lung cancer, chronic obstructive pulmonary disease, asthma, lower respiratory infections, otitis media and other conditions in both adults and children^1–3^. Recent burden-of-disease analyses estimate that SHS exposure contributed to approximately 1.3 million premature deaths worldwide in 2019, with tens of thousands of these occurring in children under 14 years ^1,2,4^. Children and adolescents are disproportionately affected, especially in low- and middle-income countries where smoke-free protections are weaker and household smoking is more common ^2,4,5^.

Global surveillance data underscore the magnitude of adolescent SHS exposure. Analysis of Global Youth Tobacco Survey (GYTS) data from more than 140 countries suggests that roughly one-third of adolescents are exposed to SHS at home and over half in public places ^4^. However, these overall estimates mask substantial regional inequalities. Systematic reviews highlight that estimates for African and other low-resource settings are often sparse, methodologically heterogeneous, or entirely absent, limiting reliable assessment of SHS-attributable disease burden ^1,5^. Serum cotinine studies further show that self-reported exposure frequently underestimates true SHS levels in children, complicating burden estimates and hampering policy evaluation^3,6^.

Sub-Saharan Africa is undergoing a tobacco transition: historically low smoking prevalence is threatened by aggressive marketing, demographic change and uneven implementation of the WHO Framework Convention on Tobacco Control (FCTC)^5,7^. Adult smoking prevalence in several countries, including Zambia, remains modest in absolute terms but is high within specific male and socio-economic subgroups ^5^. In such contexts, adolescents may experience substantial SHS exposure in homes, schools, public transport, hospitality venues, and increasingly through emerging products such as waterpipes and electronic cigarettes, which are often weakly regulated and perceived as less harmful ^8–11^. Yet, compared with evidence from Asia, Europe and North America, there are few nationally representative data describing the prevalence and determinants of adolescent SHS exposure in African settings ^1,4,5^.

Understanding the correlates of SHS exposure in adolescence is critical because this developmental period is marked by heightened vulnerability of the brain and lungs to nicotine and tobacco toxicants, as well as the consolidation of long-term health behaviors^3,12,13^. Repeated SHS exposure in childhood and adolescence impairs lung growth, increases the risk of wheeze and asthma, and contributes to cardiometabolic and blood pressure changes that may track into adulthood ^2,3,14^. At the same time, SHS often co-occurs with powerful social influences, parental and peer smoking, school and neighborhood norms, and ubiquitous tobacco marketing, that increase the likelihood of smoking initiation and progression to regular use ^4,15–18^. Identifying which socio-demographic, household, school, peer and media factors are most strongly associated with SHS exposure can therefore guide targeted interventions, from enforcing smoke-free environments to shaping mass media campaigns and youth-focused prevention programs ^4,7,16,17^.

To date, no study has comprehensively quantified the prevalence and correlates of SHS exposure among a nationally representative sample of in-school adolescents in Zambia. This study addresses that gap by estimating the burden of SHS exposure and examining its demographic, behavioral, social and environmental determinants, thereby providing essential evidence to inform FCTC implementation and adolescent health policy in the Zambian context.

## Methods

### Data management

The study used data from the 2021 Zambia Global Youth Tobacco Survey (GYTS) to conduct the analysis. The GYTS is a nationally representative, school-based, cross-sectional survey that employs a self-administered questionnaire to monitor tobacco use and related behaviors among adolescents (https://extranet.who.int/ncdsmicrodata). In Zambia, the survey was administered in grades 7–9 (primarily ages 11–17). A total of 6,499 students completed the survey (overall response rate 75.7%)^19^.

### Outcome Variables

The primary outcome was second-hand smoke (SHS) exposure. We created binary indicators for SHS exposure in four settings, based on standard GYTS questionnaire items:

- SHS at home: Any reported days in the past 7 days that someone smoked in the student’s presence inside the home (Yes vs No)^20^.
- SHS in enclosed public places: Any days in the past 7 days that someone smoked in the student’s presence inside an enclosed public place (shops, restaurants, etc., excluding home)^20^.
- SHS in outdoor public places: Any days in the past 7 days that someone smoked in the student’s presence in an outdoor public area (play grounds, sidewalks, parks, etc.).
- SHS at school: Any reported sighting of someone smoking inside the school building or on school property in the past 30 days.

Each of the above settings was coded 1 for any exposure (≥1 day or Yes) and 0 for no exposure. We also constructed a composite binary outcome (shs_any = 1) if the student was exposed in any of the four settings (and shs_any = 0 if exposed in none).

### Predictor Variables

The study examined a comprehensive set of predictor variables organized by socio-ecological level. Each predictor was coded as specified, with reference categories indicated where applicable, consistent with standard GYTS measures and previous analyses of this dataset ^19^.

- Individual-level factors: Demographics (age in years [11–17], sex [male/female], grade level [7, 8, 9]); socioeconomic status (wealth index derived from reported weekly spending money, categorized into tertiles [Low, Middle, High] as a proxy for wealth); tobacco use behaviors (ever tried cigarette smoking; current cigarette use [smoked on ≥1 days in past 30 days]; use of hand-rolled cigarettes; ever tried shisha (waterpipe); awareness of electronic cigarettes); knowledge and attitudes (belief that long-term tobacco use causes cancer; belief that long-term tobacco use leads to drug use); and future intentions (intention to use any tobacco in the next 12 months).
- Interpersonal-level factors: Family and peer influences were assessed by parental and peer smoking variables. We included whether a parent or guardian smokes in the home (Yes/No) and whether the student’s father smokes, as reported by the student. Peer influence was measured by whether any of the student’s close friends smoke tobacco. School environment factors included whether any teachers smoke in the school (teacher smoking) and whether the student has seen teachers smoking on school grounds.
- Environmental and media factors: The study assessed exposure to tobacco-related media and marketing. Media exposure was measured by core GYTS items asking how often, in the past 30 days, the student saw people using tobacco on television, in movies, or on streaming media. The study also measured exposure to tobacco advertising (e.g., seeing tobacco ads at stores or points of sale). Exposure to tobacco marketing was further captured by whether the student owned a non-tobacco item (e.g. shirt, cap) with a tobacco product brand logo. Exposure to anti-tobacco messaging was assessed through questions on whether the student had seen or heard any messages against tobacco use on TV/radio or social media.
- Policy or education factors: The study included a school-based education variable: whether the student was taught in any class about the dangers of tobacco use during the past year. (This item was coded Yes/ No.)
- Geographic factors: Region of residence was included, coded by province and grouped into Lusaka, “Rest of Country” (ROC), and Tobacco-growing regions, based on administrative region (Region Name).

All predictor variables were created according to the same definitions and coding schemes used in prior work to ensure consistency across analyses. In multivariable models, certain variables (region, age, sex, grade) were included a priori regardless of their statistical significance, following our analysis plan.

### Statistical Analysis

Data management was done in R version 4.5.2 ^21,22^. Descriptive analyses were conducted to summarise the characteristics of study participants, with results presented as frequencies and survey-weighted percentages. The weighted prevalence of secondhand smoke (SHS) exposure was estimated overall and across levels of each predictor variable. All prevalence estimates were accompanied by 95% confidence intervals (CIs) calculated using survey-adjusted methods to account for the complex sampling design of the GYTS.

Bivariate analyses were performed to examine associations between SHS exposure and individual predictor variables. Survey-weighted logistic regression models were fitted separately for each predictor, and unadjusted odds ratios (ORs) with corresponding 95% CIs were reported ^23,24^. These analyses provided an initial assessment of factors associated with SHS exposure while appropriately incorporating sampling weights, stratification, and clustering^23^. Multivariable analysis was undertaken to identify independent predictors of SHS exposure ^25^. A survey-adjusted logistic regression model including all candidate predictors was fitted to obtain the final adjusted model ^25^. Adjusted odds ratios (AORs) with 95% CIs were reported, with statistical significance assessed at the 5% level.

### Ethics

This study was conducted in accordance with internationally accepted ethical standards for research involving human participants. Access to the anonymized 2021 GYTS dataset was obtained through the World Health Organization (WHO) NCD Microdata Repository. The GYTS data are fully de-identified prior to release, ensuring that no individual respondents can be identified. As the analysis involved secondary use of existing anonymized data and required no direct contact with participants, additional ethical approval or informed consent was not required. The study therefore posed minimal risk and complied with WHO data access and research ethics requirements. Access to the raw data can be requested from the WHO through the WHO NCD Microdata Repository (http://extranet.who.int/ncdsmicrodata/index.php/catalog/957) or by contacting the WHO NCD Surveillance Team at, subject to approval and relevant data-sharing agreements. The analytical code used in this study is available from the corresponding author upon reasonable request.

## RESULTS

### Descriptive Analysis

Table 1 presents the characteristics of the study sample comprising 6,499 adolescents aged 11–17 years from three regions in Zambia: Lusaka (n = 2,027), Rest of Country (ROC; n = 2,500), and Tobacco Regions (n = 1,972). The age distribution was centered on mid-adolescence, with 25% (n = 1,589) aged 14 years and 21% (n = 1,363) aged 15 years. Adolescents aged 11 years constituted 4.1% (n = 265), while those aged 17 years accounted for 11% (n = 678). Females comprised 57% (n = 3,681) of the sample, and males 43% (n = 2,725). Students were evenly distributed across grades, with 35% in Grade 7, 32% in Grade 8, and 33% in Grade 9.

**Table 1:** Characteristics of school-going adolescents in Zambia: 2021 Global Youth Tobacco Survey.

| Characteristic | Lusaka<br>n = 2,027 | ROC<br>n = 2,500 | Tobacco<br>Regions<br>n = 1,972 | Overall<br>n = 6,499 |
| --- | --- | --- | --- | --- |
| **Socio-demographic characteristics** |  |  |  |  |
| **Age (years)** |  |  |  |  |
| 11 | 53 (2.6) | 85 (3.4) | 127 (6.5) | 265 (4.1) |
| 12 | 176 (8.7) | 166 (6.7) | 115 (5.9) | 457 (7.1) |
| 13 | 389 (19) | 388 (16) | 299 (15) | 1,076 (17) |
| 14 | 498 (25) | 608 (25) | 483 (25) | 1,589 (25) |
| 15 | 432 (21) | 547 (22) | 384 (20) | 1,363 (21) |
| 16 | 289 (14) | 422 (17) | 316 (16) | 1,027 (16) |
| 17 | 182 (9.0) | 265 (11) | 231 (12) | 678 (11) |
| **Sex** |  |  |  |  |
| Female | 1,212 (60) | 1,302 (53) | 1,167 (61) | 3,681 (57) |
| Male | 796 (40) | 1,169 (47) | 760 (39) | 2,725 (43) |
| **Grade** |  |  |  |  |
| 7 | 731 (36) | 783 (32) | 757 (39) | 2,271 (35) |
| 8 | 566 (28) | 855 (35) | 630 (33) | 2,051 (32) |
| 9 | 708 (35) | 828 (34) | 545 (28) | 2,081 (33) |
| **Wealth index** |  |  |  |  |
| Low | 1,033 (51) | 1,351 (54) | 1,032 (53) | 3,416 (53) |
| Middle | 361 (18) | 438 (18) | 338 (17) | 1,137 (18) |
| Rich | 614 (31) | 693 (28) | 586 (30) | 1,893 (29) |
| **Spending money** |  |  |  |  |
| None | 366 (18) | 562 (23) | 425 (22) | 1,353 (21) |
| Low | 667 (33) | 789 (32) | 607 (31) | 2,063 (32) |
| Medium | 585 (29) | 646 (26) | 503 (26) | 1,734 (27) |
| High | 390 (19) | 485 (20) | 421 (22) | 1,296 (20) |
| **Tobacco use behaviors** |  |  |  |  |
| Ever tried cigarette smoking | 250 (14) | 381 (20) | 452 (32) | 1,083 (21) |
| Smoked cigarettes | 140 (7.5) | 346 (16) | 433 (26) | 919 (16) |
| Tried shisha | 1,721 (86) | 2,052 (83) | 1,479 (76) | 5,252 (82) |
| Use of hand-rolled cigarette | 229 (11) | 533 (21) | 619 (31) | 1,381 (21) |
| **Household tobacco exposure** |  |  |  |  |
| Father smokes | 244 (12) | 412 (17) | 373 (19) | 1,029 (16) |
| Parent/guardian smokes in home | 244 (12) | 412 (17) | 373 (19) | 1,029 (16) |
| **School tobacco exposure** |  |  |  |  |
| Saw teachers smoking at school | 198 (9.8) | 332 (13) | 357 (18) | 887 (14) |
| Teachers smoke at school | 211 (10) | 350 (14) | 364 (18) | 925 (14) |
| **Social influences** |  |  |  |  |
| Friends smoke tobacco | 434 (22) | 624 (25) | 551 (28) | 1,609 (25) |
| **Media and advertising exposure** |  |  |  |  |
| Exposure to anti-tobacco media messages | 1,286 (64) | 1,544 (62) | 1,192 (61) | 4,022 (62) |
| See people using tobacco on TV | 1,108 (55) | 1,057 (42) | 787 (40) | 2,952 (45) |
| See tobacco advertisement | 429 (21) | 535 (21) | 405 (21) | 1,369 (21) |
| **Knowledge and perceptions** |  |  |  |  |
| Heard of electronic cigarette | 479 (24) | 626 (26) | 630 (33) | 1,735 (27) |
| Long-term tobacco use leads to drug use | 847 (42) | 983 (39) | 734 (37) | 2,564 (39) |
| Long-term tobacco use leads to cancer | 1,246 (61) | 1,413 (57) | 1,064 (54) | 3,723 (57) |
| **Attitudes and intentions** |  |  |  |  |
| Intention to use tobacco in next 5 years | 187 (9.3) | 334 (14) | 344 (18) | 865 (13) |
| Own item with tobacco product | 183 (9.0) | 314 (13) | 280 (14) | 777 (12) |
| **Education and intervention** |  |  |  |  |
| Taught about dangers of tobacco use | 1,013 (51) | 1,134 (46) | 857 (44) | 3,004 (47) |

Tobacco-related behaviors differed significantly by region (Table 1). Overall, 21% (n = 1,083) of students had ever tried cigarette smoking, with prevalence highest in Tobacco Regions (32%), followed by ROC (20%) and Lusaka (14%) (p < 0.001). Current cigarette smoking showed a similar pattern, reported by 26% of students in Tobacco Regions, 16% in ROC, and 7.5% in Lusaka (p < 0.001). Household tobacco exposure also varied regionally; parental smoking at home was reported by 16% overall, ranging from 12% in Lusaka to 19% in Tobacco Regions (p < 0.001).

School-based tobacco exposure was common. Overall, 14% of students reported seeing teachers smoking at school, and 14% reported teachers actually smoking at school, with both measures significantly higher in Tobacco Regions than in Lusaka or ROC (p < 0.001). Social influences were also prevalent, with 25% reporting friends who smoke tobacco. Exposure to anti-tobacco media messages was reported by 62% of students overall and did not differ meaningfully by region, while 47% reported having been taught about the dangers of tobacco use.

Socioeconomic indicators showed that 53% of students were classified as low wealth, 18% as middle wealth, and 29% as rich. Regarding spending money, 21% reported having none, while 32% reported low, 27% medium, and 20% high spending money.

### Prevalence of any secondhand smoke (SHS) exposure

**Table 2** presents the weighted prevalence of any secondhand smoke (SHS) exposure. Overall, 66.0 of adolescents experienced SHS exposure. Prevalence was highest among 11-year-olds (77.7) and declined through mid-adolescence before rising modestly among 17-year-olds (69.4). SHS exposure did not differ by sex and varied only slightly by grade level.

**Table 2:**
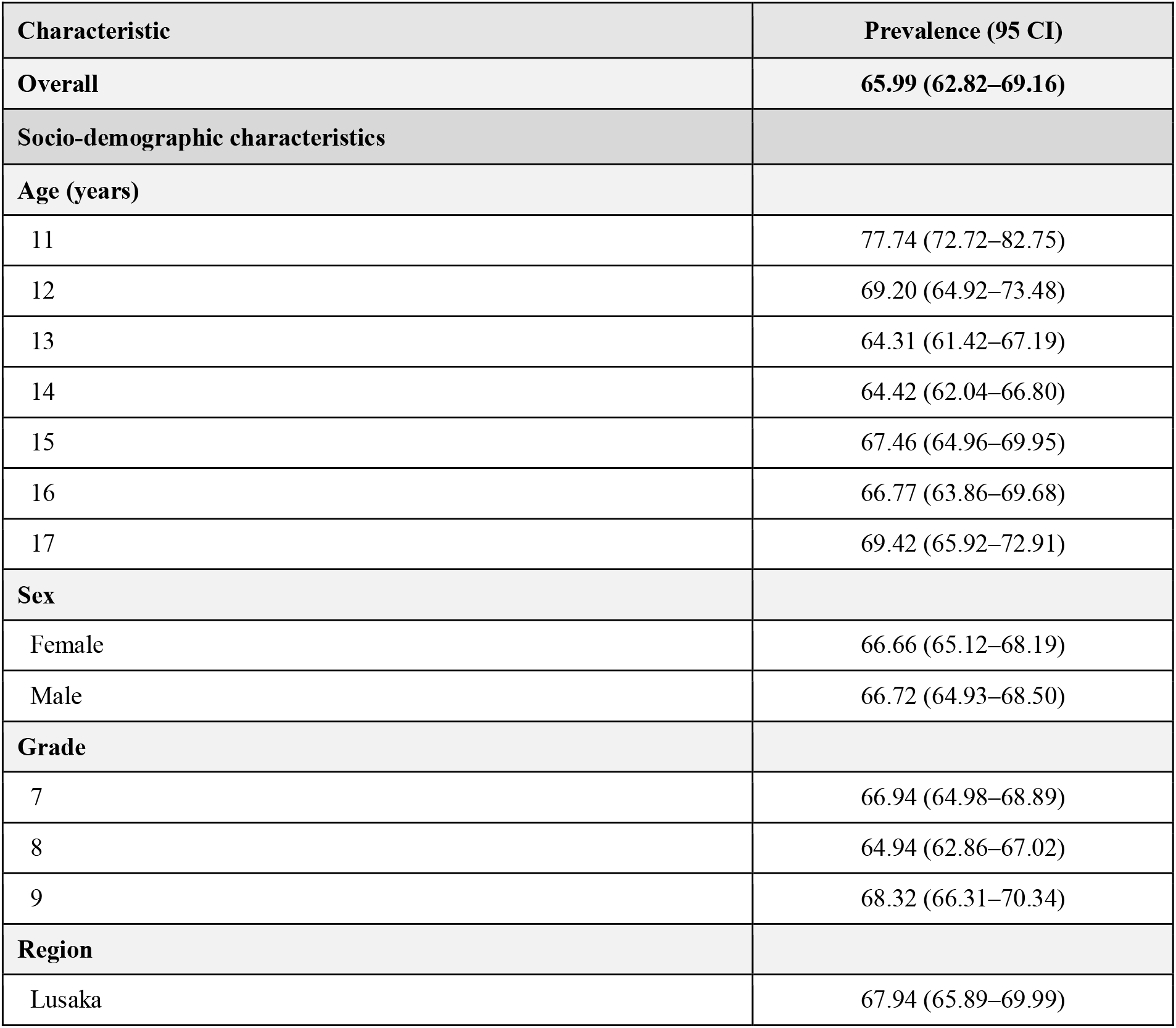
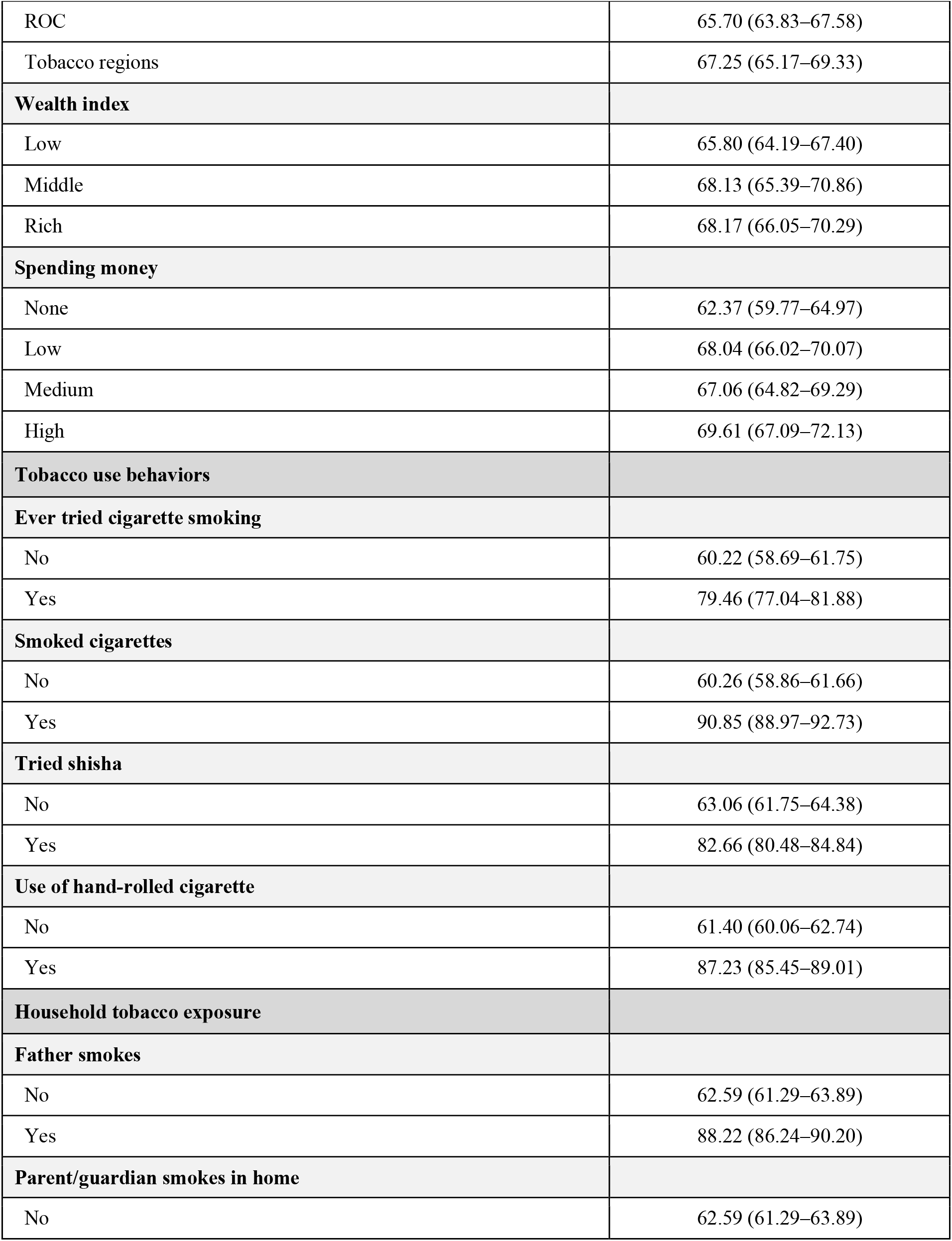
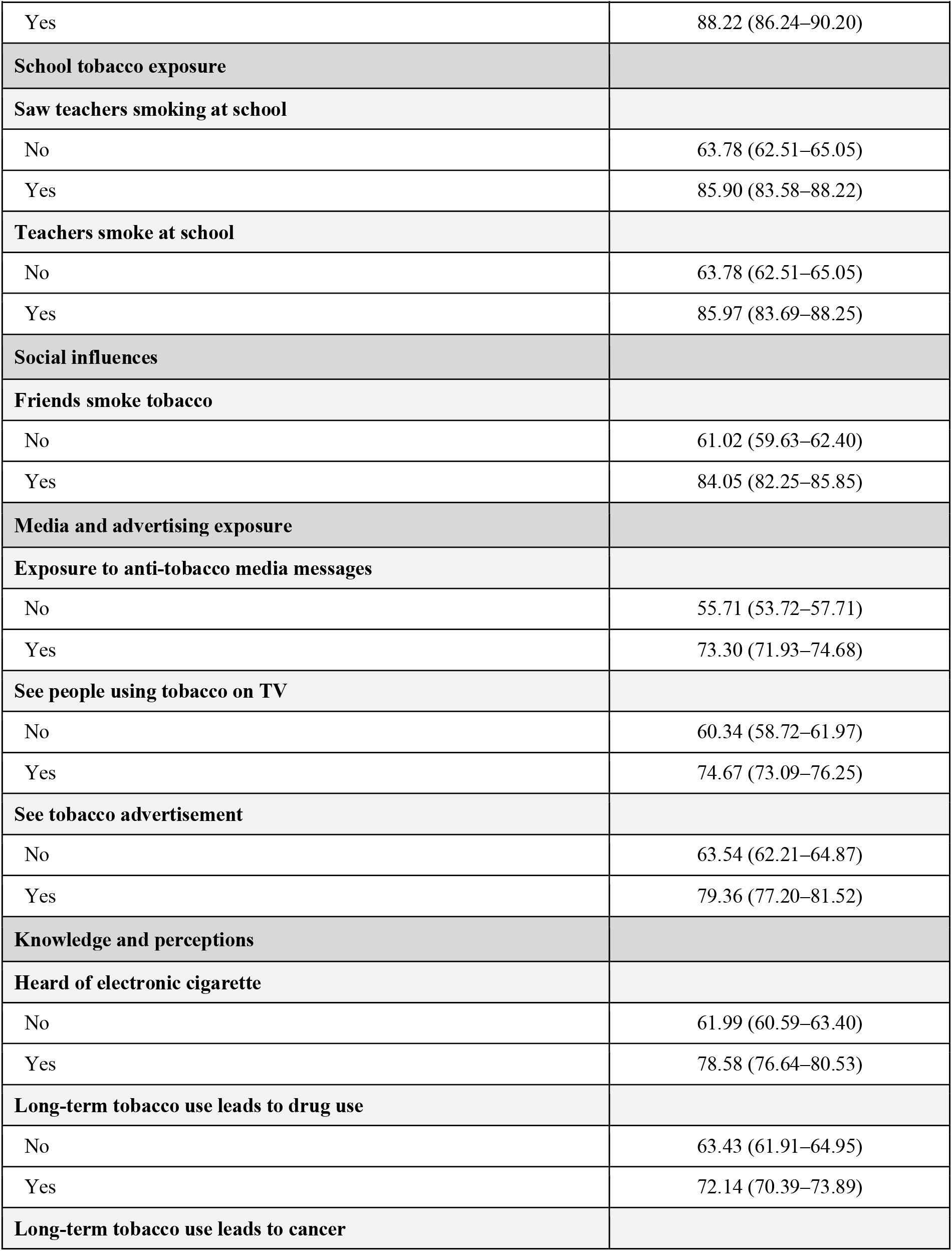
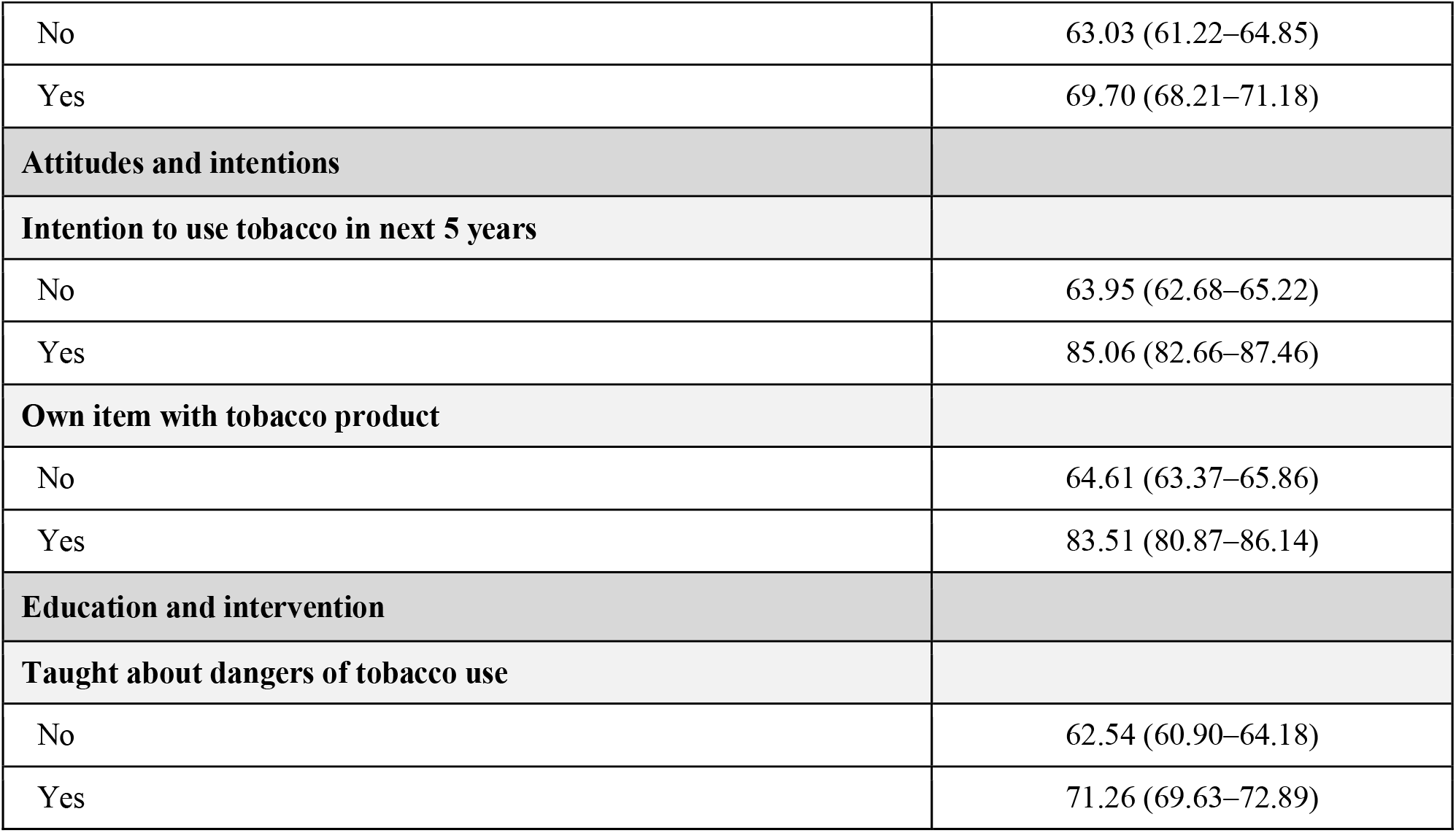
Weighted Prevalence of Exposure to Secondhand Smoking Among School-Going Adolescents in Zambia: 2021 Global Youth Tobacco Survey.

| Characteristic | Prevalence (95 CI) |
| --- | --- |
| **Overall** | **65.99 (62.82–69.16)** |
| **Socio-demographic characteristics** |  |
| **Age (years)** |  |
| 11 | 77.74 (72.72–82.75) |
| 12 | 69.20 (64.92–73.48) |
| 13 | 64.31 (61.42–67.19) |
| 14 | 64.42 (62.04–66.80) |
| 15 | 67.46 (64.96–69.95) |
| 16 | 66.77 (63.86–69.68) |
| 17 | 69.42 (65.92–72.91) |
| **Sex** |  |
| Female | 66.66 (65.12–68.19) |
| Male | 66.72 (64.93–68.50) |
| **Grade** |  |
| 7 | 66.94 (64.98–68.89) |
| 8 | 64.94 (62.86–67.02) |
| 9 | 68.32 (66.31–70.34) |
| **Region** |  |
| Lusaka | 67.94 (65.89–69.99) |
| ROC | 65.70 (63.83–67.58) |
| Tobacco regions | 67.25 (65.17–69.33) |
| **Wealth index** |  |
| Low | 65.80 (64.19–67.40) |
| Middle | 68.13 (65.39–70.86) |
| Rich | 68.17 (66.05–70.29) |
| **Spending money** |  |
| None | 62.37 (59.77–64.97) |
| Low | 68.04 (66.02–70.07) |
| Medium | 67.06 (64.82–69.29) |
| High | 69.61 (67.09–72.13) |
| **Tobacco use behaviors** |  |
| **Ever tried cigarette smoking** |  |
| No | 60.22 (58.69–61.75) |
| Yes | 79.46 (77.04–81.88) |
| **Smoked cigarettes** |  |
| No | 60.26 (58.86–61.66) |
| Yes | 90.85 (88.97–92.73) |
| **Tried shisha** |  |
| No | 63.06 (61.75–64.38) |
| Yes | 82.66 (80.48–84.84) |
| **Use of hand-rolled cigarette** |  |
| No | 61.40 (60.06–62.74) |
| Yes | 87.23 (85.45–89.01) |
| **Household tobacco exposure** |  |
| **Father smokes** |  |
| No | 62.59 (61.29–63.89) |
| Yes | 88.22 (86.24–90.20) |
| **Parent/guardian smokes in home** |  |
| No | 62.59 (61.29–63.89) |
| Yes | 88.22 (86.24–90.20) |
| **School tobacco exposure** |  |
| **Saw teachers smoking at school** |  |
| No | 63.78 (62.51–65.05) |
| Yes | 85.90 (83.58–88.22) |
| **Teachers smoke at school** |  |
| No | 63.78 (62.51–65.05) |
| Yes | 85.97 (83.69–88.25) |
| **Social influences** |  |
| **Friends smoke tobacco** |  |
| No | 61.02 (59.63–62.40) |
| Yes | 84.05 (82.25–85.85) |
| **Media and advertising exposure** |  |
| **Exposure to anti-tobacco media messages** |  |
| No | 55.71 (53.72–57.71) |
| Yes | 73.30 (71.93–74.68) |
| **See people using tobacco on TV** |  |
| No | 60.34 (58.72–61.97) |
| Yes | 74.67 (73.09–76.25) |
| **See tobacco advertisement** |  |
| No | 63.54 (62.21–64.87) |
| Yes | 79.36 (77.20–81.52) |
| **Knowledge and perceptions** |  |
| **Heard of electronic cigarette** |  |
| No | 61.99 (60.59–63.40) |
| Yes | 78.58 (76.64–80.53) |
| **Long-term tobacco use leads to drug use** |  |
| No | 63.43 (61.91–64.95) |
| Yes | 72.14 (70.39–73.89) |
| **Long-term tobacco use leads to cancer** |  |
| No | 63.03 (61.22–64.85) |
| Yes | 69.70 (68.21–71.18) |
| **Attitudes and intentions** |  |
| **Intention to use tobacco in next 5 years** |  |
| No | 63.95 (62.68–65.22) |
| Yes | 85.06 (82.66–87.46) |
| **Own item with tobacco product** |  |
| No | 64.61 (63.37–65.86) |
| Yes | 83.51 (80.87–86.14) |
| **Education and intervention** |  |
| **Taught about dangers of tobacco use** |  |
| No | 62.54 (60.90–64.18) |
| Yes | 71.26 (69.63–72.89) |

Strong gradients were observed across tobacco-related behaviors. Adolescents who had ever tried cigarette smoking reported substantially higher SHS exposure (79.5) compared with those who had not (60.2). Exposure was even higher among current cigarette smokers (90.9) and users of hand-rolled cigarettes (87.2).

Household and social environments were strongly associated with SHS exposure. Adolescents whose parent or guardian smoked in the home experienced SHS exposure at a prevalence of 88.2, compared with 62.6 among those without household smoking. Similarly, 84.1 of students with friends who smoke reported SHS exposure, compared with 61.0 among those without smoking friends. Observing teachers smoking at school was associated with SHS exposure prevalence exceeding 85.

Media exposure showed consistent associations. Adolescents who reported seeing people use tobacco on television had higher SHS exposure (74.7) than those who did not (60.3), and exposure to tobacco advertisements was associated with a prevalence of 79.4. Paradoxically, exposure to anti-tobacco media messages and being taught about the dangers of tobacco use were also associated with higher SHS exposure, likely reflecting targeted interventions in high-exposure environments. Regional differences in SHS exposure were modest, with prevalence ranging from 65.7 in ROC to 67.9 in Lusaka. Higher wealth and greater access to spending money were associated with slightly higher SHS exposure.

### Household smoking patterns

Figure 1 below summarizes household smoking patterns by region. Overall, 16.0 of adolescents reported that a parent or guardian smokes at home, and 6.7 reported daily parental smoking. Smoking by other household members was more common, reported by 21.8 overall, with daily smoking by others reported by 8.9. All household smoking indicators were more prevalent in Tobacco Regions than in Lusaka

**Figure 1.**
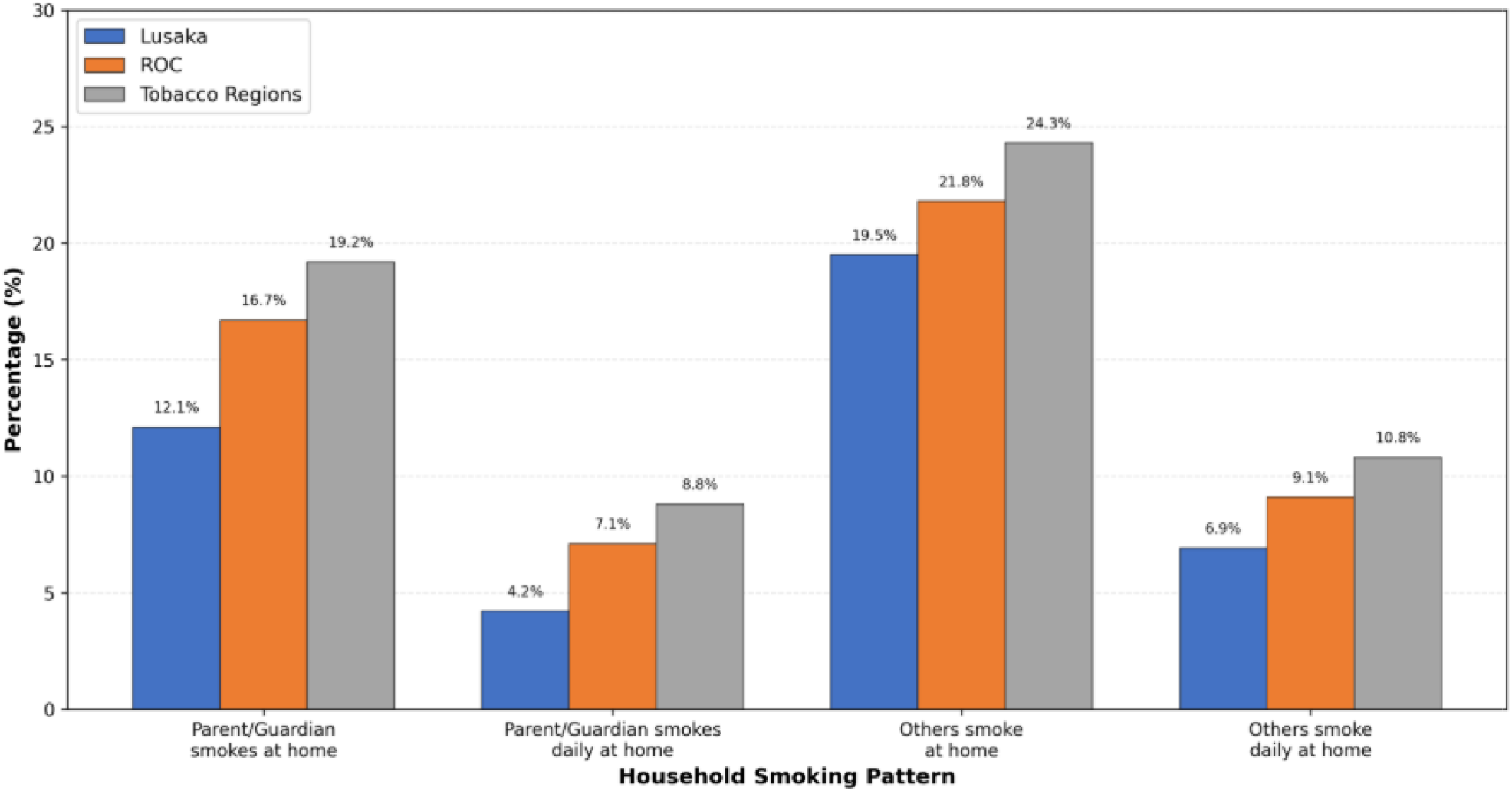
Household smoking patterns by region as reported by School-Going Adolescents in Zambia: 2021 Global Youth Tobacco Survey.

**Figure 2.**
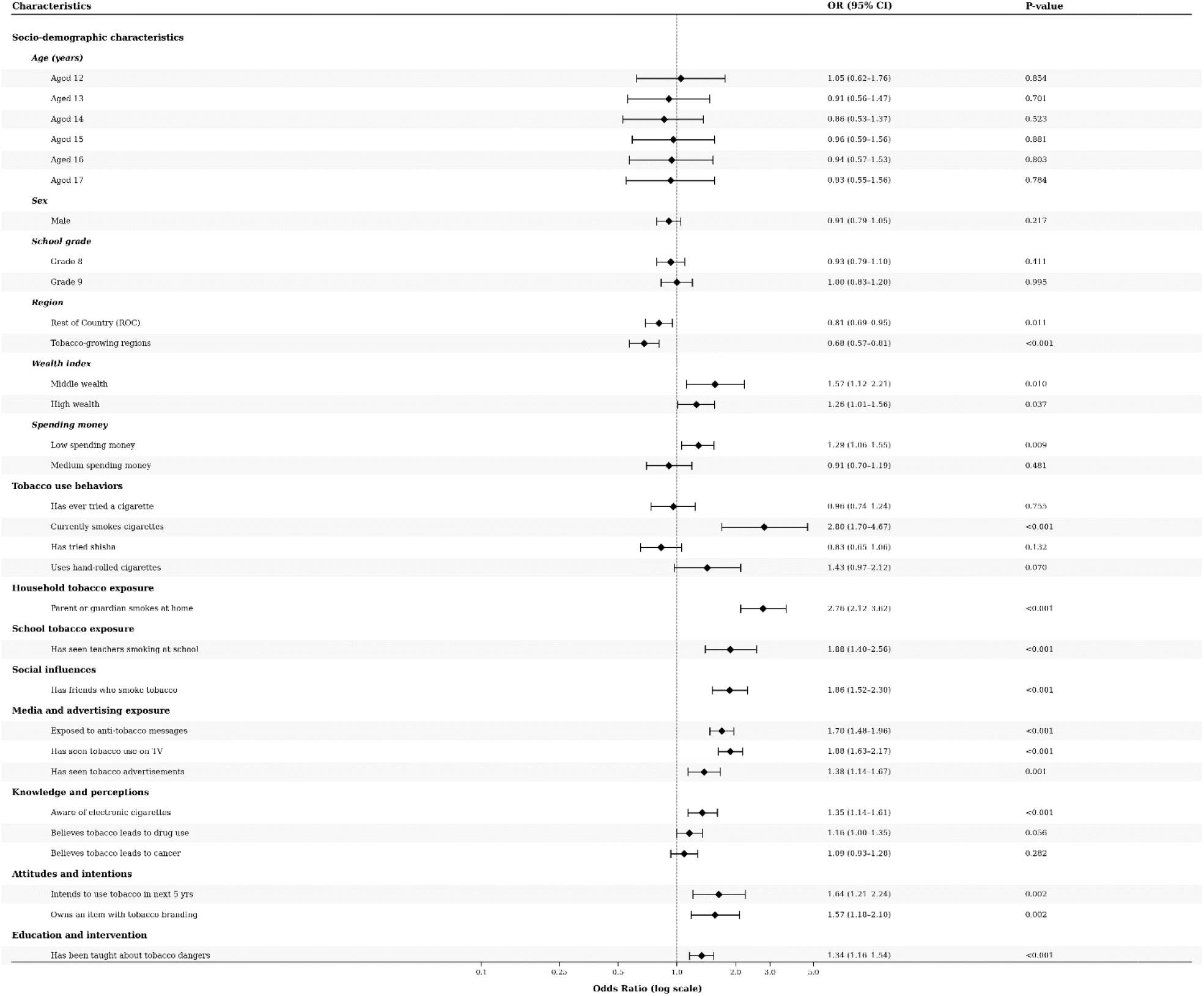
Factors Associated with exposure to secondhand smoking by school-going adolescents in Zambia: 2021 Global Youth Tobacco Survey. OR= Odds ratio; 95CI= 95 confidence interval

### Factors associated with exposure to secondhand smoking

Table 4 presents results from the multivariable logistic regression analysis examining factors associated with any secondhand smoke (SHS) exposure among adolescents. All estimates are adjusted for demographic, social, environmental, and socioeconomic factors, holding all other variables constant. Geographic region was significantly associated with SHS exposure. Compared with Lusaka, students living in the Rest of Country (ROC) had lower odds of SHS exposure (OR = 0.81, 95 CI: 0.69–0.95, p = 0.011). Adolescents in Tobacco Regions also had reduced odds compared with Lusaka (OR = 0.68, 95 CI: 0.57–0.81, p < 0.001). Age was not independently associated with SHS exposure after adjustment. Compared with 11-year-olds, 12-year-olds had slightly higher odds (OR = 1.05, 95 CI: 0.62–1.76, p = 0.854), while adolescents aged 13–17 years generally showed lower odds, ranging from OR = 0.91 among 13-year-olds (95 CI: 0.56–1.47, p = 0.701) to OR = 0.86 among 14-year-olds (95 CI: 0.53–1.37, p = 0.523). However, none of these associations were statistically significant, and all confidence intervals included one, suggesting that the observed age differences may be due to chance rather than true age-related effects.

Sex was also not associated with SHS exposure. Males had similar odds compared with females (OR = 0.91, 95 CI: 0.79–1.05, p = 0.217). Grade level showed no meaningful association, with Grade 8 students having slightly reduced odds compared with Grade 7 (OR = 0.93, 95 CI: 0.79–1.10, p = 0.411), and Grade 9 students showing no difference (OR = 1.00, 95 CI: 0.83–1.20, p = 0.995).Exposure to anti-tobacco interventions was positively associated with SHS exposure. Adolescents who reported exposure to anti-tobacco media messages had higher odds of SHS exposure (OR = 1.70, 95 CI: 1.48–1.96, p < 0.001). Similarly, those who had been taught about the dangers of tobacco use had increased odds (OR = 1.34, 95 CI: 1.16–1.54, p < 0.001). These findings likely reflect greater exposure to prevention efforts in environments where SHS exposure is already common. Social and household environments showed the strongest associations with SHS exposure. Adolescents living with a parent or guardian who smokes had nearly three times higher odds of SHS exposure (OR = 2.76, 95 CI: 2.12–3.62, p < 0.001). Having friends who smoke tobacco increased the odds by almost two times (OR = 1.86, 95 CI: 1.52–2.30, p < 0.001). Seeing teachers smoking at school was also associated with increased odds of exposure (OR = 1.88, 95 CI: 1.40–2.56, p < 0.001).

Media and commercial exposures were important predictors. Adolescents who reported seeing people using tobacco on television had almost twice the odds of SHS exposure (OR = 1.88, 95 CI: 1.63–2.17, p < 0.001). Exposure to tobacco advertisements was also associated with higher odds (OR = 1.38, 95 CI: 1.14–1.67, p = 0.001). Personal tobacco-related behaviours and perceptions were significantly associated with SHS exposure. Adolescents who had smoked cigarettes had nearly three times higher odds of exposure (OR = 2.80, 95 CI: 1.70–4.67, p < 0.001). Awareness of electronic cigarettes increased the odds (OR = 1.35, 95 CI: 1.14–1.61, p < 0.001), while ownership of tobacco-branded items was associated with higher odds (OR = 1.57, 95 CI: 1.18–2.10, p = 0.002). Adolescents who intended to use tobacco in the next five years also had increased odds of current SHS exposure (OR = 1.64, 95 CI: 1.21– 2.24, p = 0.002). In contrast, having ever tried cigarette smoking was not independently associated with SHS exposure (OR = 0.96, 95 CI: 0.74–1.24, p = 0.755). Tobacco-related knowledge showed mixed associations. Belief that long-term tobacco use leads to drug use showed a borderline association with SHS exposure (OR = 1.16, 95 CI: 1.00–1.35, p = 0.056), while belief that tobacco use leads to cancer was not associated with SHS exposure (OR = 1.09, 95 CI: 0.93–1.28, p = 0.282).

Socioeconomic factors showed varying patterns. Compared with adolescents with no spending money, those with low spending money had higher odds of SHS exposure (OR = 1.29, 95 CI: 1.06–1.55, p = 0.009), while medium spending money showed no association (OR = 0.91, 95 CI: 0.70–1.19, p = 0.481). Compared with adolescents in the low wealth category, those in the middle wealth group had higher odds of SHS exposure (OR = 1.57, 95 CI: 1.12–2.21, p = 0.010), and those in the high wealth group also showed increased odds (OR = 1.26, 95 CI: 1.01–1.56, p = 0.037). Some variables were not independently estimated due to collinearity or lack of statistical significance. Trying shisha (OR = 0.83, 95 CI: 0.65–1.06, p = 0.132) and use of hand-rolled cigarettes (OR = 1.43, 95 CI: 0.97–2.12, p = 0.070) were not significant.

## DISCUSSION

The prevalence of secondhand smoke (SHS) exposure among Zambian adolescents stands out as one of the highest globally, with 66 reporting recent exposure. This figure is not only higher than the pooled estimate for African adolescents, which ranges from 12.5 to 45, but also surpasses rates reported in Nigeria (43), West Africa (45), and Malaysia (41–53)^26–31^. It is also above the global average of 55.9 among adolescents in 68 low- and middle-income countries ^29^, though still below the extreme 97 reported in The Gambia ^32^. Such variation in prevalence across countries can be attributed to differences in adult and youth smoking rates, enforcement and coverage of smoke-free laws, urbanization, and social norms regarding tobacco use at home, school, and public places^29,32,33^.

In Zambia, SHS exposure was highest among 11-year-olds (77.7), declined through mid-adolescence, then rose again by age 17 (69.4). However, after adjustment for confounders, age was not independently associated with SHS exposure, contrasting with studies from Cambodia, Mongolia, Taiwan, and the US that found increasing age correlated with higher SHS exposure ^6,33–35^. Sex was also not a significant factor; males and females had similar odds of SHS exposure. This aligns with some Malaysian studies but contrasts with US data where girls were more susceptible to SHS ^36,37^. The lack of sex difference in Zambia may reflect local patterns of tobacco use and prevailing social norms.

Strong gradients were observed across tobacco-related behaviors. Adolescents who had ever tried cigarette smoking reported much higher SHS exposure (79.5) compared to those who had not (60.2). Exposure was even higher among current cigarette smokers (90.9) and users of hand-rolled cigarettes (87.2). These findings are consistent with studies from Malaysia, China, South Africa, Japan, and the US showing that adolescent smokers are far more likely to be exposed to SHS, likely due to clustering within peer groups who share smoking behaviors ^29,30,34,38,39^. Smokers tend to befriend other smokers as they share a common behavior; more than 90 of adolescents tend to smoke together in groups ^40^, which increases their collective exposure to SHS.

Household and social environments were strongly associated with SHS exposure. Adolescents whose parent or guardian smoked at home experienced an SHS prevalence of 88.2, compared to 62.6 among those without household smoking, a pattern echoed across Africa, Asia, Latin America, and high-income countries ^27,28,33,41–43^. Similarly, having friends who smoke increased SHS exposure to 84.1 versus 61 for those without such friends; observing teachers smoking at school was associated with prevalence exceeding 85. These results are consistent with international evidence that parental, peer, and teacher smoking are among the strongest predictors of youth SHS exposure ^27,28,41^. In many settings, including The Gambia and West Africa, parental education level, living arrangements, home smoking rules, and being sent to purchase cigarettes all significantly increase risk of adolescent SHS exposure ^32^.

Media influences also play a significant role. Zambian adolescents who saw people use tobacco on television had higher SHS exposure (74.7) than those who did not (60.3), while exposure to tobacco advertisements was associated with a prevalence of 79.4. Unexpectedly, seeing anti-tobacco media messages or being taught about tobacco dangers was also linked to higher SHS exposure, likely reflecting targeted interventions in high-exposure environments or greater recall among frequently exposed youths ^44^. Similar “paradoxical” associations have been documented in West African GYTS data and pooled African analyses ^45^.

Regional differences within Zambia were modest: prevalence ranged from 65.7 in Rest of Country to 67.9 in Lusaka; students outside Lusaka had lower odds of SHS exposure after adjustment, a pattern seen elsewhere where urbanization increases risk ^40^. Higher wealth status and greater access to spending money were associated with slightly higher SHS exposure, a complex pattern also observed in Kuwait and Malaysia where both low-SES home exposures and high-SES social exposures contribute^30,39^.

Multivariable analysis confirmed that living with a parent or guardian who smokes nearly tripled odds of SHS exposure; having friends who smoke nearly doubled odds; seeing teachers smoke at school increased odds by nearly double, media or commercial exposures remained significant predictors; personal tobacco use strongly predicted higher odds; while age, sex, grade level showed no independent association. The Zambian findings fit within a broader literature showing that where enforcement of smoke-free legislation is weak, and adult or peer smoking remains common, the majority of adolescents are exposed to SHS. The strong influence of family or peer or teacher smoking has been repeatedly demonstrated across Africa, Asia, Europe, North America, Latin America, and is further reinforced by biomarker-based studies using cotinine levels that confirm self-reported exposures often underestimate true prevalence ^6^. The paradoxical association between anti-tobacco education or media messages and higher reported SHS has also been observed elsewhere, often interpreted as reverse causation or targeted messaging in high-risk settings ^44^.

Socioeconomic disparities persist, children from lower-income families or rural areas are often more exposed due to crowded housing conditions or less stringent enforcement of home bans ^33,46^, while wealthier youths may encounter more public or social exposures as seen in Hong Kong’s rising neighbor-sourced household smoke trends ^46^. Thirdhand smoke, residual nicotine on surfaces, has emerged as an additional concern for children even when active smoking is absent at home or school environments remain contaminated from prior use ^35^.

Health consequences are significant: beyond respiratory illness and asthma aggravation well-documented globally ^2^, recent research links adolescent SHS exposure to depressive symptoms ^47^, negative emotions such as loneliness or anxiety ^48^, sleep disturbances ^49^, myopia progression ^42^, metabolic syndrome risk factors including dyslipidemia or insulin resistance ^50^, elevated blood pressure or hypertension risk ^38^, under-five mortality in sub-Saharan Africa ^30^, academic performance deficits ^51^, increased susceptibility to future tobacco use initiation, even among never-smokers, and clustering with other risky behaviors such as alcohol or drug use or e-cigarette experimentation via secondhand vape exposures now rising worldwide ^52^.

Measurement approaches vary. While most studies rely on self-report surveys like GYTS/GSHS for cross-national comparability despite recall bias concerns^29^, biomarker-based assessments using serum cotinine provide objective confirmation but remain less feasible for large-scale surveillance^6^. Both methods consistently show that comprehensive policy interventions, smoke-free laws covering homes or cars or public spaces combined with parental cessation support, are needed for meaningful reductions^2,53^.

In conclusion, exposure to secondhand smoke among adolescents in Zambia remains high. This exposure is largely shaped by smoking in the home, peer influence, and smoking by teachers at school, despite existing smoke-free policies. Media exposure and tobacco marketing further contribute to continued exposure, while socioeconomic conditions also play an important role. Reducing secondhand smoke exposure among adolescents will require action across multiple settings, including stronger enforcement of smoke-free school and public space policies, greater support for parents and caregivers to quit smoking, and interventions that address peer norms and media influences. Anti-tobacco education should be better targeted to reach adolescents in high-exposure environments and across all socioeconomic groups. Without coordinated, multi-level interventions supported by ongoing surveillance, secondhand smoke exposure among adolescents is likely to remain a major public health concern in Zambia.

## Abbreviations

SSA: Sub-Saharan Africa
SHS: Secondhand Smoke
GYTS: Global Youth Tobacco Survey
WHO: World Health Organization
FCTC: Framework Convention on Tobacco Control
LMICs: Low- and Middle-Income Countries
OR: Odds Ratio
CI: Confidence Interval
ROC: Rest of Country

## Acknowledgements

The authors would like to thank the World Health Organization, the Ministry of Health Zambia, participating schools, students, and survey administrators for their contribution to the 2021 Zambia Global Youth Tobacco Survey (GYTS), which made this analysis possible.

## Author contributions

WN led the conceptualisation of the study, conducted the statistical analysis, and drafted the manuscript. SM, AC, and SH contributed to study conceptualisation and critically reviewed the analysis and manuscript. CZ provided policy-relevant insights and reviewed the manuscript. All authors reviewed and approved the final version of the paper.

## Data availability

The data analyzed in this study are from the 2021 Zambia Global Youth Tobacco Survey (GYTS) and are publicly available through the World Health Organization NCD Microdata Repository. Access to the data is subject to World Health Organization data access approval procedures.

## Declarations

### Ethics approval and consent to participate

The 2021 Zambia Global Youth Tobacco Survey (GYTS) was conducted in accordance with the ethical principles outlined in the Declaration of Helsinki. Ethical approval was obtained from the relevant national ethics authorities in Zambia, with technical oversight from the World Health Organization. Written informed consent was obtained from all participating students prior to data collection.

This study is a secondary analysis of de-identified, publicly available GYTS data accessed through an approved World Health Organization data request. The analysis involved no direct contact with participants and did not require additional ethical approval.

### Consent for publication

Not applicable. This manuscript does not contain any individual-level identifiable data.

